# The Prediction for the Outbreak of COVID-19 for 15 States in USA by Using Turning Phase Concepts as of April 10, 2020

**DOI:** 10.1101/2020.04.13.20064048

**Authors:** George Xianzhi Yuan, Lan Di, Yudi Gu, Guoqi Qian, Xiaosong Qian

## Abstract

Based on a new concept called “Turning Period”, the goal of this report is to show how we can conduct the prediction for the outlook in the different stages for the battle with outbreak of COVID-19 currently in US, in particular, to identify when each of top 15 states in USA (basically on their populations) is going to enter into the stage that the outbreak of COVID-19 is under the control by the criteria such as daily change of new patients is less than 10% smoothly. Indeed, based on the data of April 10, 2020 with the numerical analysis, we are able to classify 15 states of US into the following four different categories for the Prevention and Control of Infectious Diseases Today and the main conclusion are:

**First**, staring **around April 14, 20202**, three states which are **Washington State, Louisiana** and **Indiana** are entering the stage that the outbreak of COVID-19 is under the control, which means daily change of new patients is less than 10% and the gamma is less than zero in general.

**Second**, staring **<u>around</u> <u>April</u> <u>15</u>, 20202**, two states which are **New Jersey, and New Yor**k are entering the stage that the outbreak of COVID-19 is under the control, which means daily change of new patients is less than 10% and the gamma is less than zero in general.

**Third**, staring **<u>around</u> <u>April</u> <u>16</u>, 20202**, seven states which are **California, Florida, Georgia (GA), Illinois, Maryland, Indiana, Michigan**, and **Pennsylvania** are entering the stage that the outbreak of COVID-19 is under the control, which means daily change of new patients is less than 10% and the gamma is less than zero in general.

**Fourth**, staring **<u>around</u> <u>April</u> <u>17</u>, 20202**, three states which are **Texas, Massachusetts**, and **Connecticut** are entering the stage that the outbreak of COVID-19 is under the control, which means daily change of new patients is less than 10% and the gamma is less than zero in general.

Finally, we want to reinforce that emergency risk management is always associated with the implementation of an emergency plan. The identification of the Turning Time Period is key to emergency planning as it provides a timeline for effective actions and solutions to combat a pandemic by reducing as much unexpected risk as soon as possible.

## 1 The Background and Related Literature

Infectious disease epidemics always present challenges to human society, threatening the safety of human life and causing social upheaval and economic losses. In recent years, novel virus outbreaks have been increasing worldwide, from the 2003 SARS-CoV, the H1N1 influenza A virus in 2009, the MERS-CoV in 2012, the Ebola virus in 2015, the Chai virus in 2016, the H5N7 avian influenza virus in 2017, to the recent coronavirus (COVID-19) that emerged at the end of 2019 (in Wuhan, China). These outbreaks have brought great loss to human life, disrupted population processes, and negatively impacted global development.

The current coronavirus disease 2019 (COVID-19) outbreak began in Wuhan City (Hubei province of China) in later December 2019 and quickly spread to other cities in China in a matter of days. It was announced as a public health emergency of international concern by the World Health Organization (WHO) on January 30, 2020. Predicting the development of the development of the outbreak as early and as reliably as possible is critical for action to prevent its spread with necessary implementation of emergency response plan.

Furthermore, the current COVID-19 outbreak is a particularly urgent public health event. Apart from the challenges the virus presented to China’s health system, it has spread rapidly to other regions and countries worldwide. We currently do not have a unanimous metric of estimating when the global situation of the virus will be under control.

In this study, we like to discuss how to establish a general framework for the prediction of the critical so-called “Turning Period” which would play a very important role in assisting better plans for the time frame of emergence plans, in particular for associated looking forward planning such as the battle with the current pandemics of COVID-19 worldwide.

As applications, we will show which level (situation) of 13 countries in Europe (consisting of Germany, France, Italy, Spain, Belgium, Austria, Switzerland, Norway, Netherlands, Portugal, Sweden, UK, and Russia) are in fighting with the outbreak of OCOVID-19 as of April 9, 2020.

Indeed, our study also indicates that the implementation of the emergency program in the practice associated with the “Isolation Control Program (or, say, Wuhan Quarantine Program (see Begley [36])) “since January23, 2020 by China in national level may be a good experiences by other countries and regions to take a lesson.

During almost last century, in the study and modelling mechanics of infectious diseases, the traditional model called “SEIR” denoted for “infectious disease dynamics susceptible–exposed– infectious–resistant” and its various (deterministic) versions have been introduced and been very popular in analyzing and predicting the development of an epidemic (see Liu et al. [1], Murray [2], Wu et al. [3-4], Prem et al. [5], Li et al.[6], Lin et al.[7], Kuniya [8], Roosa et al. [9] and references wherein). The SEIR models the flows of people between four states: susceptible state variable “S”, exposed variable “E”, infected variable “I”, and resistant variable “R”. Each of those variables represents the number of people in those groups. Take COVID-19 as one example, assume that the average number of exposed cases that are generated by one infected person of COVID-19, this number could be regarded as the so-called “**basic reproduction number**” (which is indeed the expected number of cases directly generated by one case in a population where all individuals are susceptible to infection), the study on the basic reproduction number, related features for the globally stable endemic and disease-free equilibria and thresholds is always the main stream for people from the academic research community to the practice in the subject of epidemic disease spread behavior and related social issues.

In particular, a great deal and effort have been done for the study on the process and evolution of the limits of the Basic Reproduction Number and similar thresholds in predicting global dynamics of epidemics. In particular, since the occurrence of COVID-19 later December, 2019 in Wuhan, the study on the impact such as how serious the outbreak of infectious disease to the society, and to prediction how many people would be infected to become infectious, and so on, have been attracted by a large number of scholars with reports, see Cao et al. [10-11], Cowling and Leung [12], Hermanowicz [13], Li et al. [14], Guan et al. [15] and reference wherein.

On the other hand, modelling the situation of COVID-19 and effects of different containment strategies in China with dynamic differential equations and parameters estimation have also been paid a lot attention by a number of scholars, e.g., see Gu et al. [17], Hu et al. [18], Zhao et al. [18], Yan et al. [20], Wang et al. [21], Tang et al. [22], Huang et al [23], Cui and Hu [24] and related references wherein.

In particular, Professor Murray [2] leads his IHME COVID-19 health service utilization forecasting team to work on the estimates of predicted health service utilization and deaths due to COVID-19 by day for the next 4 months for each state in the US. Their objective is to determine the extent and timing of deaths and excess demand for hospital services due to COVID-19 in the US (also, see the study of Kuniya [8] on Japan, Murray [2] on USA, Wu et al. [3-4], Prem et al. [5] on China).

It seems that almost all of them still follow the way to pay the attention mainly on modelling or forecasting the behavior of spread for epidemic disease directly related to those infected who also become infectious, i.e., the variable “I” of SEIR model. No study pays the attention on the study how to establish a general framework for the prediction of the critical turning period for the spread of pandemic diseases (e.g., the outbreak of COVID-19) in general.

The object of this paper is to fill in this gap as we do believe that it is so important to study the general dynamics for the outbreak of COVID-19 in each country or regions, which faces one simple expectation that how to find in which time period the battle with COVID-19 will be under controlled ? By a simple fact that for any spread of infectious disease, we know that in general it is impossible to find or identify the exact turning point (or critical point) for a big pandemic (which means the behavior of the spread of COVID-19 virus is under control) due to various dynamics and associated uncertainty! But if using the idea to think of the a time period (or say, time interval) instead of an exact time point to identify the true change for the behavior of spread for epidemic disease such as in terms of the number of infectious people have significantly be reduced, plus the population of the exposed ⋹ are also true under the control to reach to certain low level, then it seems possible for us to identify different phases and stages for the mechanics of the outbreak of infectious disease incorporating with some useful tools such as the iSEIR one we introduced in [28] (see also [38]).

As suggested by the title of the paper, the goal is to conduct the prediction for the outbreak of COVId-19 in 15 states in US by using a new concept called Turning Phase using information as of April 9, 2020, we will first discuss how to build the framework for the prediction of the Critical Turning Period for Outbreak of pandemics (e.g., the spread of COVID-19) based on the application of our iSEIR Model. It expects that the concept for the prediction of the critical turning period would provide us a better way to prepare the emergency plan for the prevention and control of COVID-19 in the practice, such as working on the estimates of predicted health service utilization and deaths due to COVID-19 by day for the next 4 months for each state in the US (see [3] for more information), countries and regions worldwide.

Actually, as reported by Tan [35] (see also Yuan et al. [38]) by assessing the performance of prediction by using the iSEIR model for the timeline of the spread’s mechanics of COVID-19 in Wuhan on dates of Feb.6 and Feb.10, 2020 by using the concept of “Turning Time Period (Time Period)” to forecast the time frame for the control of the epidemic outbreak measured by a reduction in the number of people infected, it shows that our iSEIR model (an extension of the SEIR model) works very well to accurately predicted that “the COVID-19 situation in China would peak around mid-to late February as early as February 7, 2020”. This review also shows that the identification of the Turning Time Period is the key to have a successful implementation for emergency plan as it provides a timeline for effective actions and solutions to combat a pandemic by reducing as much unexpected risk as soon as possible.

This paper consists of 4 sections as follows.

## 2 The Challenges faced by the Emergency Mechanism of Epidemic Prevention and Control of Infectious Diseases Worldwide Today

The idea of the key “**SEIR** Epidemic Model” can be traced back to Dr. Ronald Ross who received the Nobel Prize for Physiology or Medicine in 1902 for his work on malaria which laid the foundation for combatting the epidemic disease (see [28], and also [38]). In 1927, Kermack and McKendrick formulated a simple deterministic model called SIR to describe the dynamic mechanism for directly transmitted viral or bacterial agent in a closed population (see [26]). Since then, scholars have contributed and advanced this field; a significant milestone in the study of Epidemics was the publication of “The Mathematical Theory of Infectious Diseases” in 1957 by Bailey (see [27]). Of these, the famous SEIR model, a core subject in Epidemic discipline (see [3]), like mentioned above, is the basis for describing the mechanism for the spread of infectious diseases, and has been used in a number of research projects and related applications. In the SEIR model, the “S” state refers to the susceptible group (or ignorants) who are susceptible to disease but have not been infected yet; the “E” state refers to the exposed group who are infected but are not infectious yet (or lurkers); the “I” state refers to those infected who also become infectious; and the “R” state refers to those who have recovered from the infection (through treatment or natural recovery) who may or may no longer be infectious, or those who have passed away.

### 2.1 The Challenges faced by the Emergency Mechanism of Epidemic Prevention and Control

Infectious diseases have always been a major challenge to human society, threatening the safety of human life and causing social upheaval and economic losses. Every scenario of an epidemic outbreak due to a novel infectious disease carries a similar set of challenges: the unknown nature of the new pathogen/strain, a lack of immediate effective treatment and vaccine, and an ill-prepared public health infrastructure to accommodate the surge in potential patients and need for testing. Public health policies that could alleviate and help prevent the impact and scale of an outbreak require significant and massive governmental and societal implementation of emergency planning and intervention strategies. At present, I want to focus the objective of this paper towards **three key issues** in approaching these outbreaks:

1. **How do we establish a spatiotemporal model for the infectious diseases’ outbreak:** in order describe the mechanics of the spread of infectious diseases?
2. **How do we conduct numerical simulation and risk prediction indicators**: in order to conduct numerical simulation based on the real scenes, which can be used to provide an outlook and planning schedule associated with a key period known as the “**Turning Phase**” during the spread of infectious diseases?
3. **How do we carry out effective predictive analysis on the epidemic situation of infectious diseases on an ongoing basis:** in order to cooperate with dynamic management, support public health emergency plans/services, and support community responses by establishing a coherent bigdata method for data fusion from different sources with different structure.

In combatting these outbreaks, the immediate implementation of an emergency response mechanism delays an epidemic’s peak which affords us more time to control the epidemic by reducing the number of infections in a concentrated period of time. Thus, a successful emergency plan lengthens the “Turning Phase” (or say, “Turning time Period” (see [29], [30] and also [34]), the time interval between T0 and T1. Effective ways of flattening the curve include intervention actions such distancing or isolation programs (e.g., quarantine program (see [33]-[35] and [36]).

Thus, a major challenge faced by our current responses to epidemic prevention and infectious disease control is finding a way to predict the critical time period “**Turning Period**” or “**Turning Phase**” when implementing an emergency plan. Knowing this timeline is critical to combat the outbreak of an epidemic or infectious disease.

In this section, we discuss the framework for predicting the “**Turning Phase**” using our **iSEIR Model** introduced in [28] which was used to successfully predict the “Turning Phase” for the outbreak of epidemic COVID-19 virus from January 2020 to early of March 2020 in China using only data from Feb. 10, 2020. Here we give a brief introduction for our **iSEIR mode**, which stands for the name “**individual Susceptible-Exposed-Infective-Removed**”. The description of iSEIR is given by **Appendix 1** below (see also Yuan et al. [38] in details).

## 3 The Framework for the Prediction of the Critical “Turning Phase” based on our iSEIR Model for the Emergency Implementation Response in an Epidemic Infectious Disease Outbreak

In this section, we discuss the framework for predicting the “**Turning Phase**” using our **iSEIR Model** introduced in [28] which was used to successfully predict the “Turning Phase” for the outbreak of epidemic COVID-19 virus from January 2020 to early of March 2020 in China using only data from Feb. 10, 2020. As previously discussed, in the battle for epidemic prevention and control of infectious disease outbreak, it is crucial to implement effective prevention measures in the early stages of an outbreak. Furthermore, identifying the beginning and ending points of the time interval which forms the “**Turning Time Period** (**Time Period**)” lets us know how long to expect to implement emergency protocols to effectively flatten the curve. It is possible to make this predictive analysis of the “Turning Time Period” using the iSEIR model through the “**Supersaturation Phenomenon**” elaborated below (see [28]).

### 3.1 The Concept of “Turning Time Period (Turing Period)” for Turning Phase

While the exact turning point (or critical point) for any infectious disease spread cannot be precisely determined due to various dynamics and associated uncertainty, by borrowing the variables of “**Delta**” and “**Gamma**” practiced in financial risk management (see Hull [37]) it is possible to identify the upper and lower limits using, for example, the current tolerable degrees from the change in new (confirmed) patients daily. These indicators then allow us to identify the different phases and stages of an infectious disease outbreak through the iSEIR model, which we elaborate in Part A and Part B. Through this method, it is possible to calculate the critical time period of the “Turning Time Phase (Turning Phase).”

#### Part A: To Identify Different Time Phases of Epidemic Infectious Diseases Spread

We propose the identification of three general three phases (time periods) for the emergency response of Epidemic Infectious Diseases Spread paired with medical response actions as elaborated below:

1. **The First Phase**: The initial starting stage corresponds to the initial occurrence and prepare for possible emergence plan of a new virus which may or may not transform into a new epidemic.
2. **The Second Phase**: For our consideration, this is the most important phase which is the so-called “**First Half-Time Phase**” otherwise known as the “**Turning Phase**” (or “**Turning Period**”) which starts with the beginning of a possible outbreak and includes the delayed epidemic peak by implementation of emergency planning to control the disease spread. The First Half Time phase involves Delta and Gamma indicators (elaborated more in Part B below) to measure the daily change in number of new patients (i.e., the indicator “Delta”), and the rate of the daily change in number of new patients (i.e., the indicator “Gamma”). As shown in Figure 1 above, the time interval from T0 to T1 is our Turning Phase (Turning Period), and also makes the end of the “First Half Time Phase” for an epidemic infectious diseases spread.

**Figure 1:**
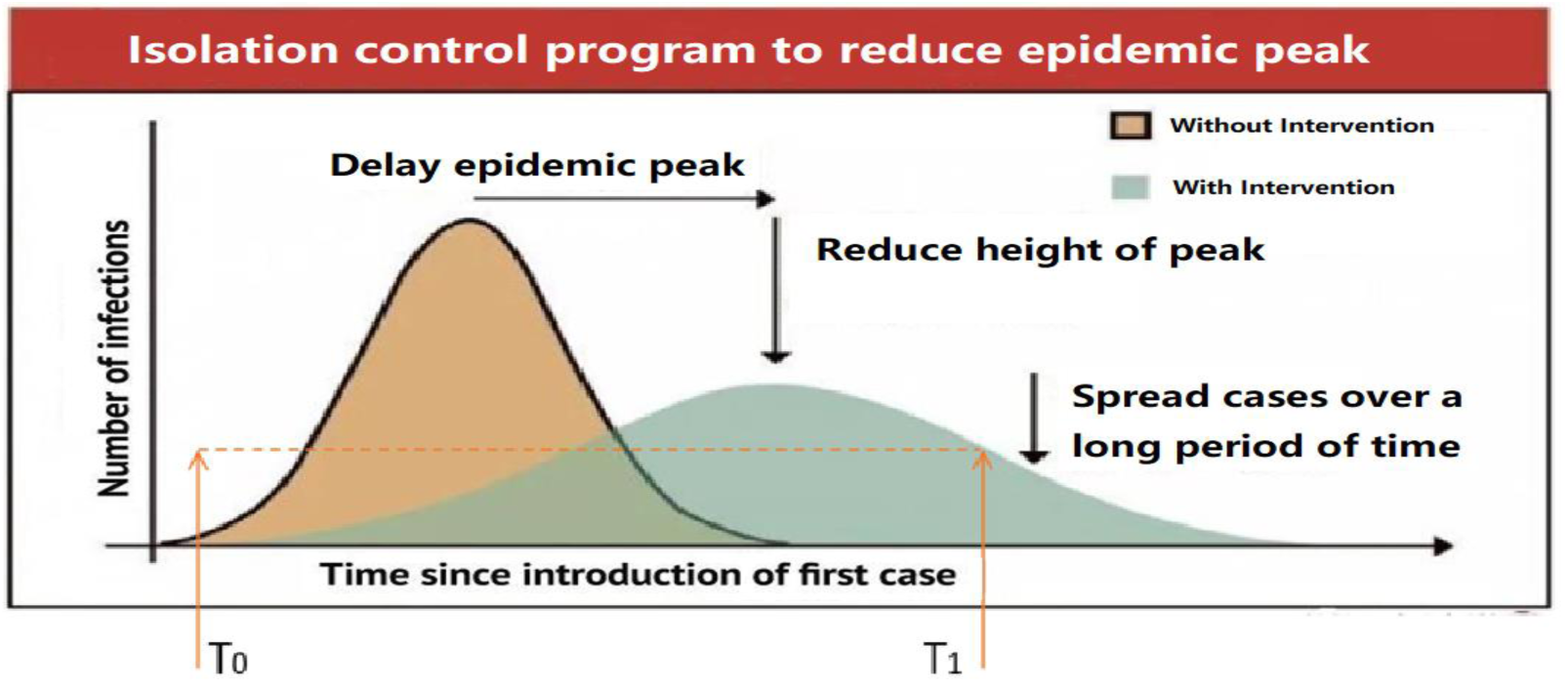

**Figure 2:**
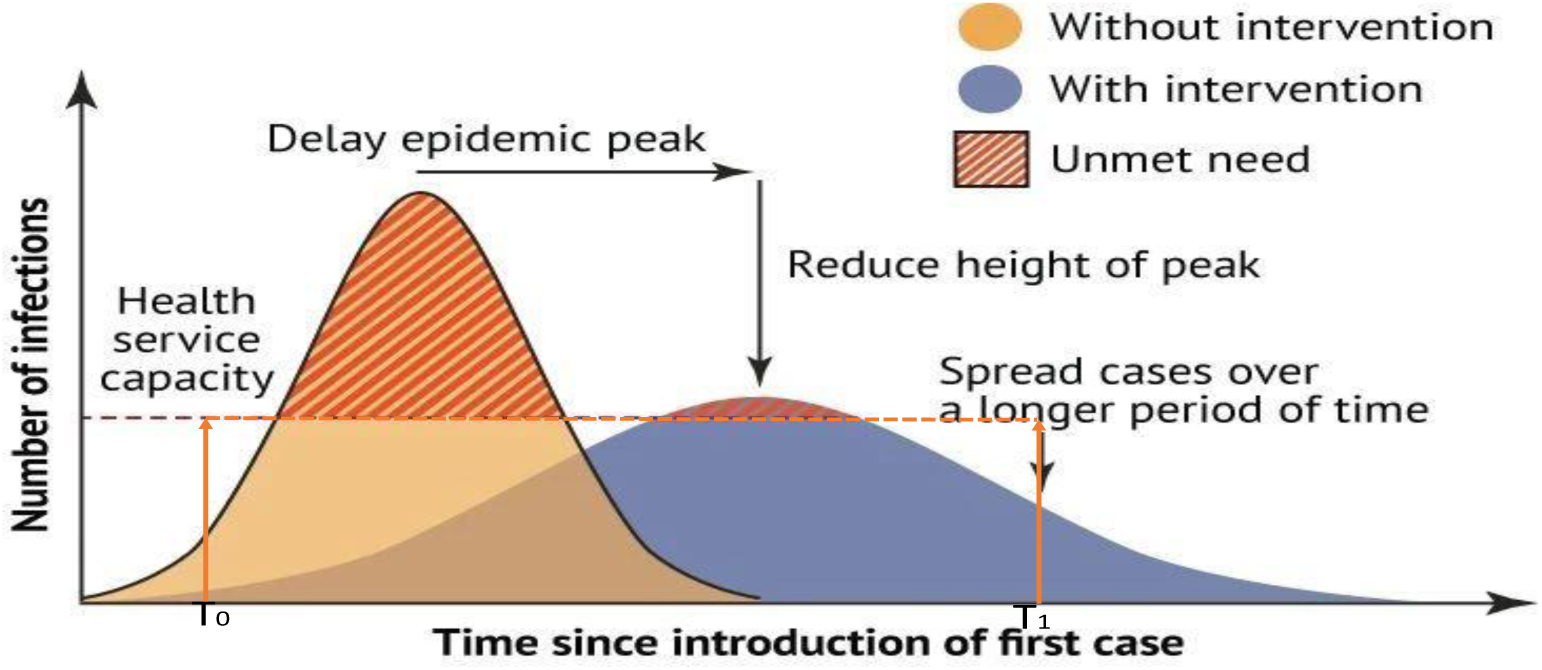
3. **The Third Phase**: During this stage, the epidemic infectious disease spread enters the do-called “**Second Half -Time Phase**”, which means the epidemic peak is gone and the rate of spread is under greater control. This is measured in a continuously decreasing rate of new infections per day, and ultimately leads to any but not exclusively of the following scenarios: a): the disease completely disappears; b): an effective vaccine/treatment is introduced; and c): the strain could also disappear and reappear cyclically in seasons, or other reasons.

Of the three phases, the most important time period to identify is the beginning and the ending time points of the **First-Half Phase** (known as the “**Turning Period**”). This phase is crucial to controlling the outbreak and spread of an epidemic infectious disease after the first case of occurrence.

Thus, being able to identify the “**First Half-Phase**” is crucial for the reliable prediction of the “**Turning Period**” (or “**Turning Phase**”) as the ending time point of the Turing Period will allow us to predict when the outbreak of the infectious disease is under the control by the level we may settle (incorporating with ability and capability in the practice).

The next challenge to address is how to identify or predict this Turning Period: To determine the Turning Period, we look to the occurrence of the so-called “Supersaturation Phenomenon” (elaborated below) based on our iSEIR model (see [28]-[31] and also the report by [34]) by running the simulation for the four control variables S(t), E(t), I(t) and R(t) in the iSEIR model (see [28]). These variables are functions of time “t” under the probability framework of individuals involved in the epidemic disease’s spread. Our prediction can be achieved once we observe the so-called “**supersaturation phenomenon**”, **the moment where the future value range of T1 observes both E(t) and I(t) decrease**. We determine this by simulating an iSEIR model that incorporates data from the initial daily disease spread (further explained by Part B below) (see also Figure 3 for both E(t) and I(t)) going down).

**Figure 3:**
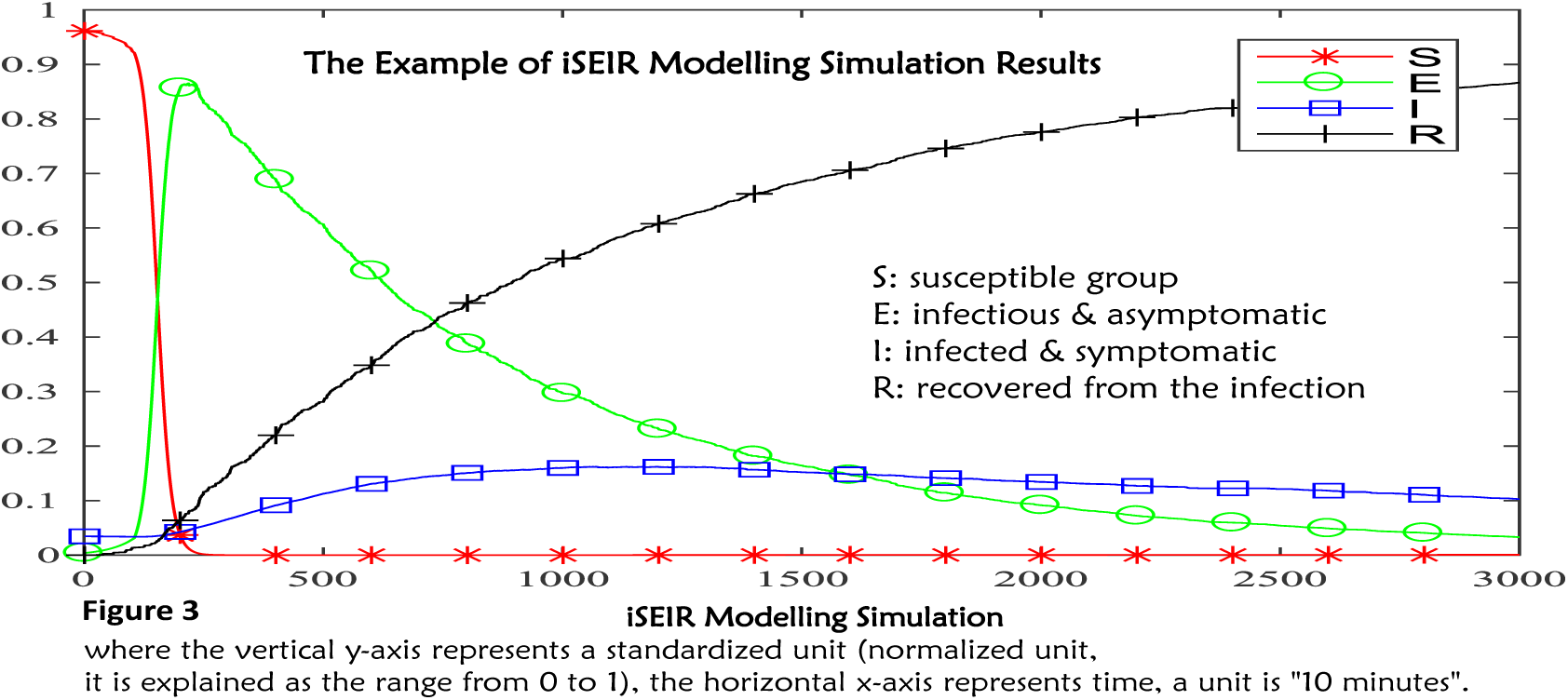

#### Part B: The Prediction of the Turning Period by using iSEIR Model Associated with Delta and Gamma Risk Indicators

When COVID-19 first emerged, we suggested using Delta and Gamma indicators to measure the daily change of new patients and the change rate of the daily change for new patients. These variables allowed us to identify the beginning point of the time interval for the Turning Period within China. For example, to identify the starting point T0 of the Turning Period, the level we considered for Delta is settled as “**no greater than 10% daily in last 6 consecutive days”**, and the average Gamma is “**no larger than 0% in last 6 consecutive days”** (we will use this criteria to define the meaning of “outbreak of COVID-19 is under the control” used below for the study of 15 states’ situation based on official data released by each state on April 10 2020).

**Next, to predict the future ending time point T1 of the Turning Period, which measures when** the epidemic disease spread is under our control, we run numerical simulations of our iSEIR model as shown by Figure 3. T1 is determined to be the point at which both control variables E(t) and I(t) drop.

The combination of Part A and Part B allowed us to reliably predict the Turning Period of COVID-19 since its first case in Wuhan (China) in late December 2019. Using only the available data released by the National Health Commission of China from Feb. 10, 2020, we correctly assessed that “**COVID-19 peak around mid-to later February, and entering in the Second Half Period around Feb. 20, 2020**.” (see our report [31] and confirmed by WHO’s report [34]).

To further understand our predictive simulation, we briefly describe the new idea of our iSEIR model and the related “**Supersaturation Phenomenon**”.

### 3.2 Our iSEIR Model as a new tool to model for “Supersaturation Phenomenon”

Almost all key models such as SEIR and related mathematical tools that exist to model the mechanics of epidemic disease spread are established under deterministic frameworks which assume that all individual behaviors and patterns are uniform (i.e., all behaviors of individuals are homogeneous), but this is not true as each individual’s behavior of infecting or being infected is different. In order to have a better way of describing the dynamics of “**Spreading behavior**” in multiplex network at an individual level to community to population levels, we introduced the so-called **iSEIR model (individual Susceptible-Exposed-Infective-Removed)** which operates under a probability perspective for each individual (see reference [28]) around two years ago as an extension of the classic SEIR one. This **iSEIR** model allows us to conduct simulations from the individual level located on the nodes of different community networks by incorporating its uncertainty with the probability to conduct random scenarios study with consideration of the corresponding multiplex networks. The behavior distribution of S, E, I and R can also be numerically simulated from the **iSEIR** model with properly specified values of parameters on population scales (in the change of percentage), population density, and transfer rate and so to have the simulation results given by Figure 3 above.

Thus, the simulation results based on the iSEIR model suggest that the intensity and extensiveness of the spread of the disease can be lowered by external intervention under a so-called “**supersaturation phenomenon**.” This phenomenon occurs when at some point in the future (denoted by T1) when both of the variables “E(t)” and “I(t)” drop in value and do not increase anymore as shown around the x-axis value of 1500 units in Figure 3.

At this value, both variables “E(t)” and “I(t)” are at the “supersaturation phenomenon.” Thus, the “supersaturation phenomenon” in the iSEIR model allows us to predict the turning period for the outbreak of epidemic diseases spread in the application.

We also like to share with readers that the general study on the prediction of the “peak period” for the outbreak of COVID-19” in China for early February 2020 has been given by Yuan et al. [38], and also see the report given by Tan [35].

### 4 The Prediction for the 15 States in USA in fighting with Outbreak of COVID-19 by Using Turning Phase Concepts as of April 10, 2020

We believe that our iSEIR model is a reliable predictor for the Turning Period of the struggle against COVID-19 through the concepts of “Turning Phase” and “Supersaturation Phenomenon”. In this section, we show how we conduct Prediction for the 15 states^2^ in US in fighting with Outbreak of COVID-19 by Using Turning Phase Concepts as of April 10, 2020 based on the framework discussed above. The following are the summary of the official data released to World Health Organization (WHO) on April 10, 2020 for 15 states in US:

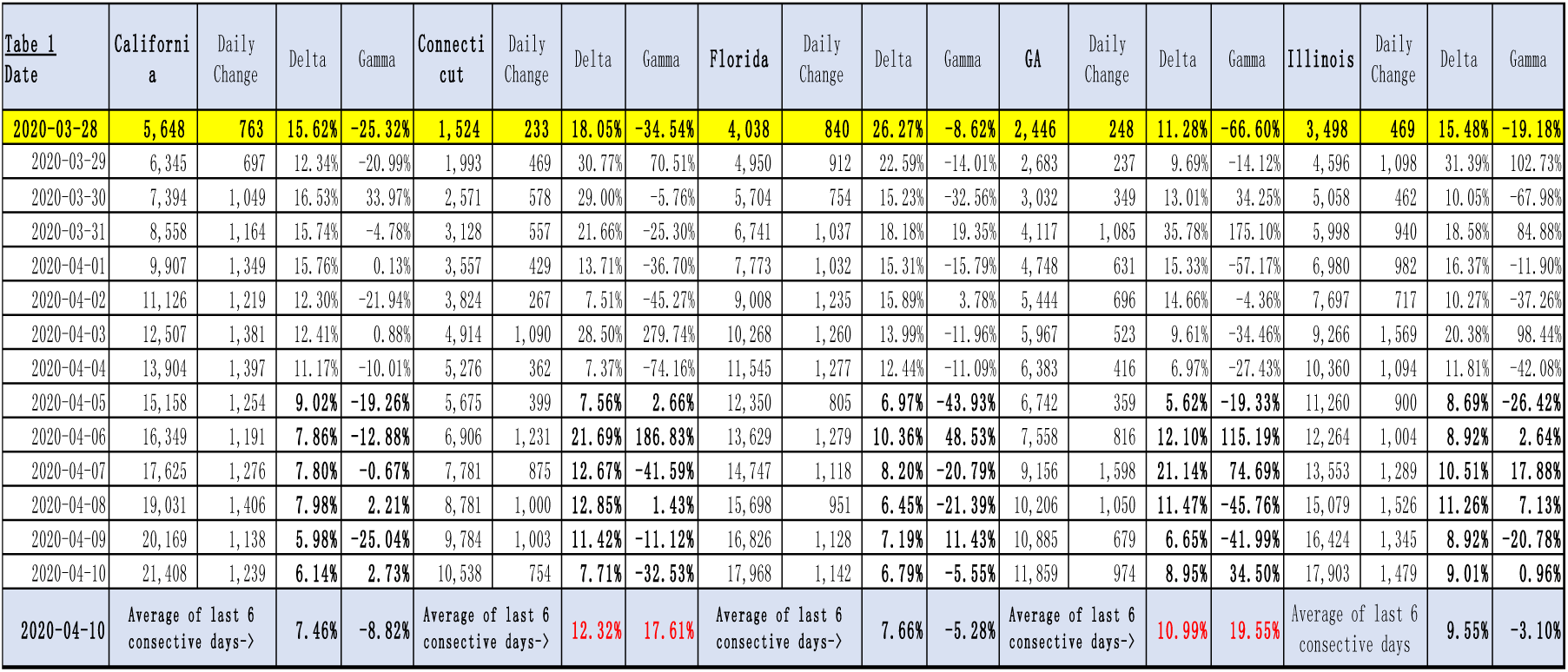

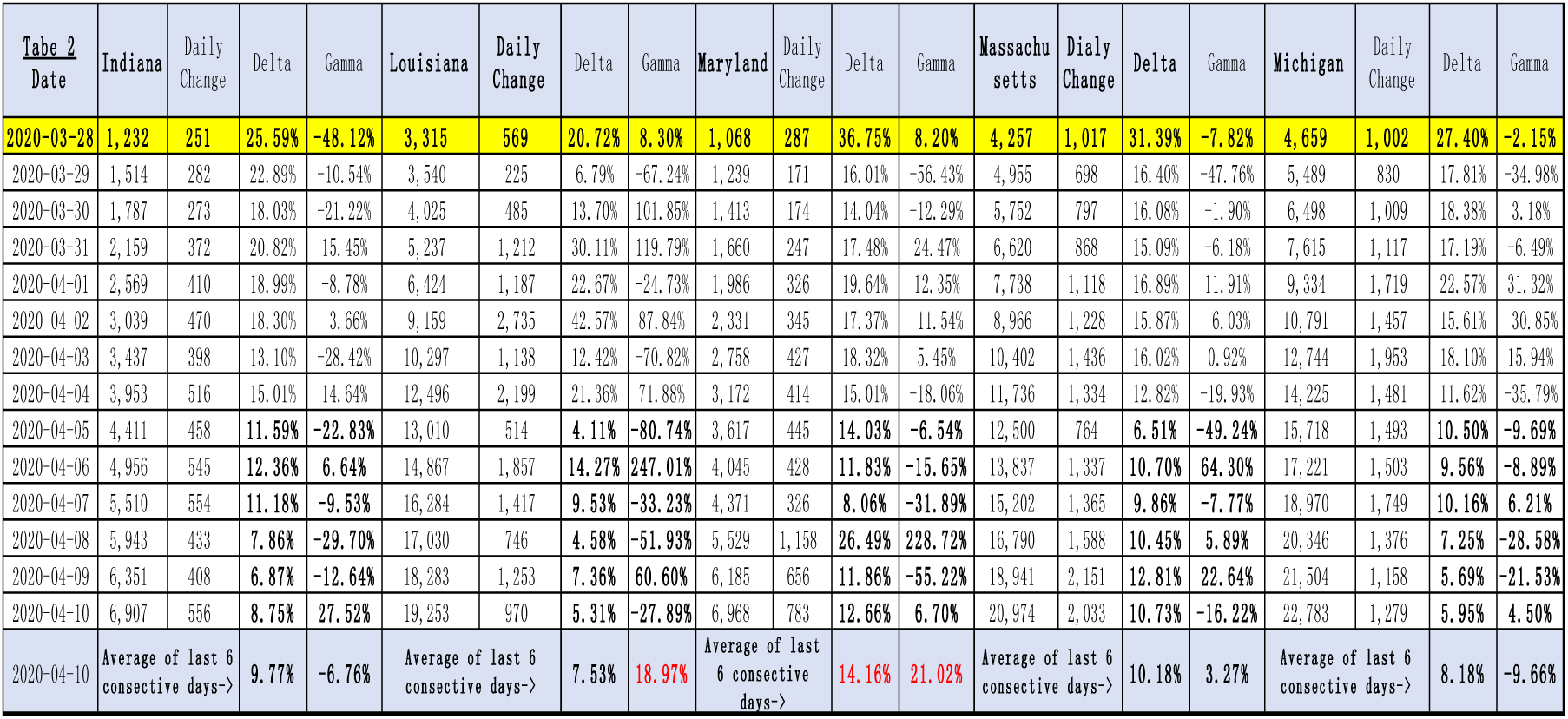

and

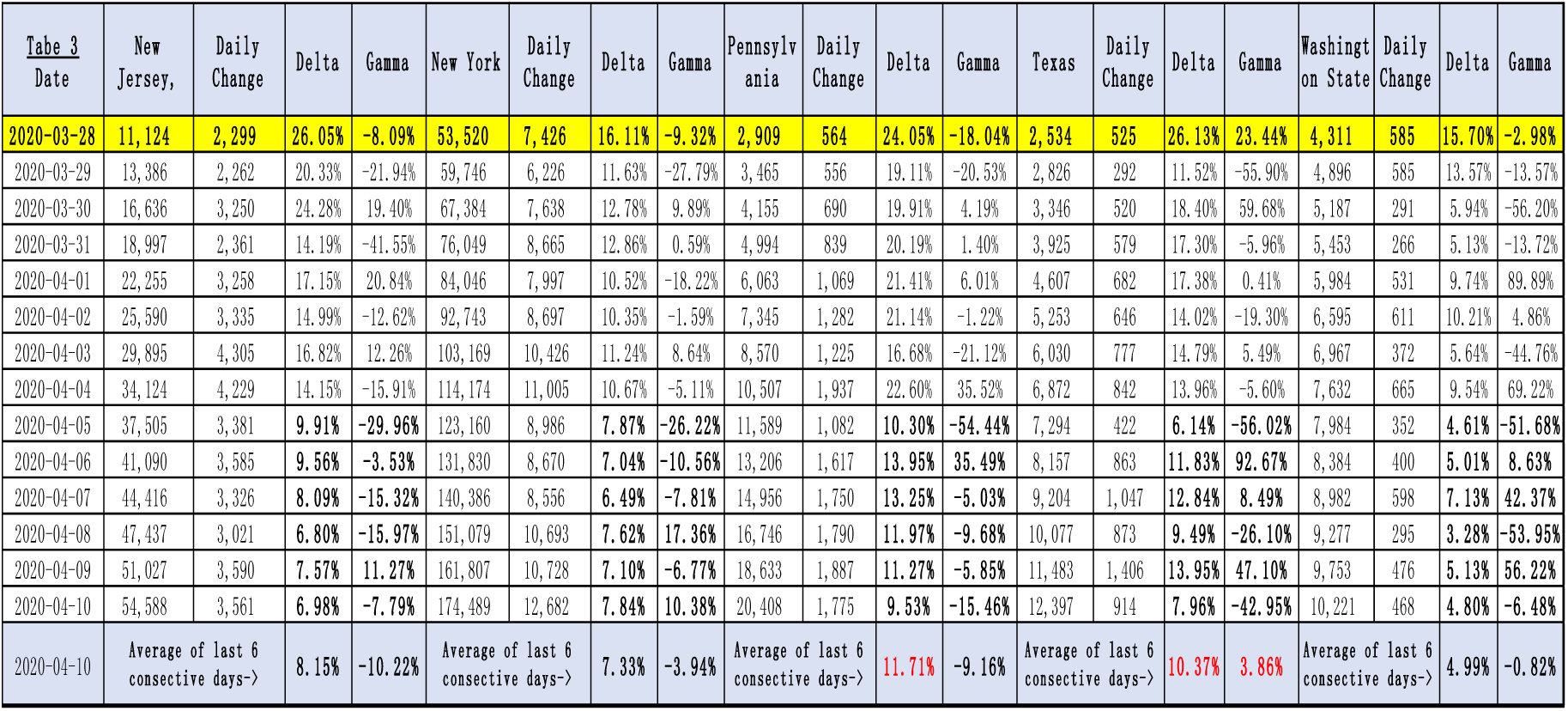

For each of 15 states, we also include their delta and gamma based on the daily change of new patients since March 29 to April 10, 2020. It seem that based on the last 6 consecutive days back from April 10, 2020, we have that:

1. The delta and gamma for most states are less than 10%, and 0% respectively, except that for Connecticut, its average delta is 12.32%, its average gamma is 17.61%; for GA, its average gamma is 19.55%; for Louisiana, its average gamma is 18.97%; for Maryland, its average delta is 14.16%, and its average gamma is 21.02%;
2. On the other hand, by following the framework established in Section 3 above, and using the criteria to identify the beginning of Turning period (phase) for “Delta” being “**less than 10% for 6 consecutive days**”, plus the average of Gamma is “**less than 0% in 6 consecutive days**”, then the above three tables show the 7 states now entering the stage of “Turning Time Period (Turning Phase)” for the control of outbreak for COVID-19 are: California, Florida, Illinois, Michigan, New Jersey, New York, and Washington state.

However, as framework established in Session 3 above, the best way to identify if the State is truly entering the Turing Phase is to see if the ending points of Turing Phase is able to be observed through simulations for the so-called “**Supersaturation Phenomenon**” with data as of April 10, 2020 in applying iSEIR models.

The following Table4 is the information for the current situations of 15 States in US as of April 10, 2020 in fighting with COVID-19 as a part inputs:

| Table 4<br>2020/4/10 | California | Connecticut | Florida | Georgia | Illinois | Indiana | Louisiana | Maryland | Massachusetts | Michigan | New Jersey | New York | Pennsylvania | Texas | Washington State |
| --- | --- | --- | --- | --- | --- | --- | --- | --- | --- | --- | --- | --- | --- | --- | --- |
| Confirmed cases (cumulative) | 21,408 | 10,538 | 17,968 | 11,859 | 17,903 | 6,907 | 19,253 | 6,968 | 20,974 | 22,783 | 54,588 | 174,489 | 20,408 | 12,397 | 10,221 |
| Death cases (cumulative) | 599 | 448 | 419 | 425 | 597 | 300 | 755 | 171 | 599 | 1,281 | 1,932 | 7,887 | 449 | 248 | 484 |
| Mortality rate | 2.80% | 4.25% | 2.33% | 3.58% | 3.33% | 4.34% | 3.92% | 2.45% | 2.86% | 5.62% | 3.54% | 4.52% | 2.20% | 2.00% | 4.74% |
| Population (million) | 39.7 | 5.8 | 21.3 | 10.5 | 12.7 | 6.7 | 4.7 | 6.9 | 6.9 | 10 | 9 | 19 | 12.8 | 28.7 | 7.5 |
| Recovery Rate | 3.6% | 0.0% | 0.0% | 0.0% | 1.2% | 0.8% | 0.0% | 4.1% | 3.5% | 0.4% | 0.1% | 9.5% | 0.8% | 13.8% | 11.2% |

Plus Incorporating data specified by **Appendix 2** below and considering each State’s situation with the implementation of “distance program” in three different levels in terms of the “distribution density” parameter ρ *by taking values being* 0.4 (within a kind of closing distance), 0.2(meaning a reasonable distance), and 0.05 (which means the people are almost in the state being separated, or isolated), then we have the following simulations results given by Figures 4, 5, 6 and 7 below and they will help us to conduct some key conclusions

**Figure 4:**
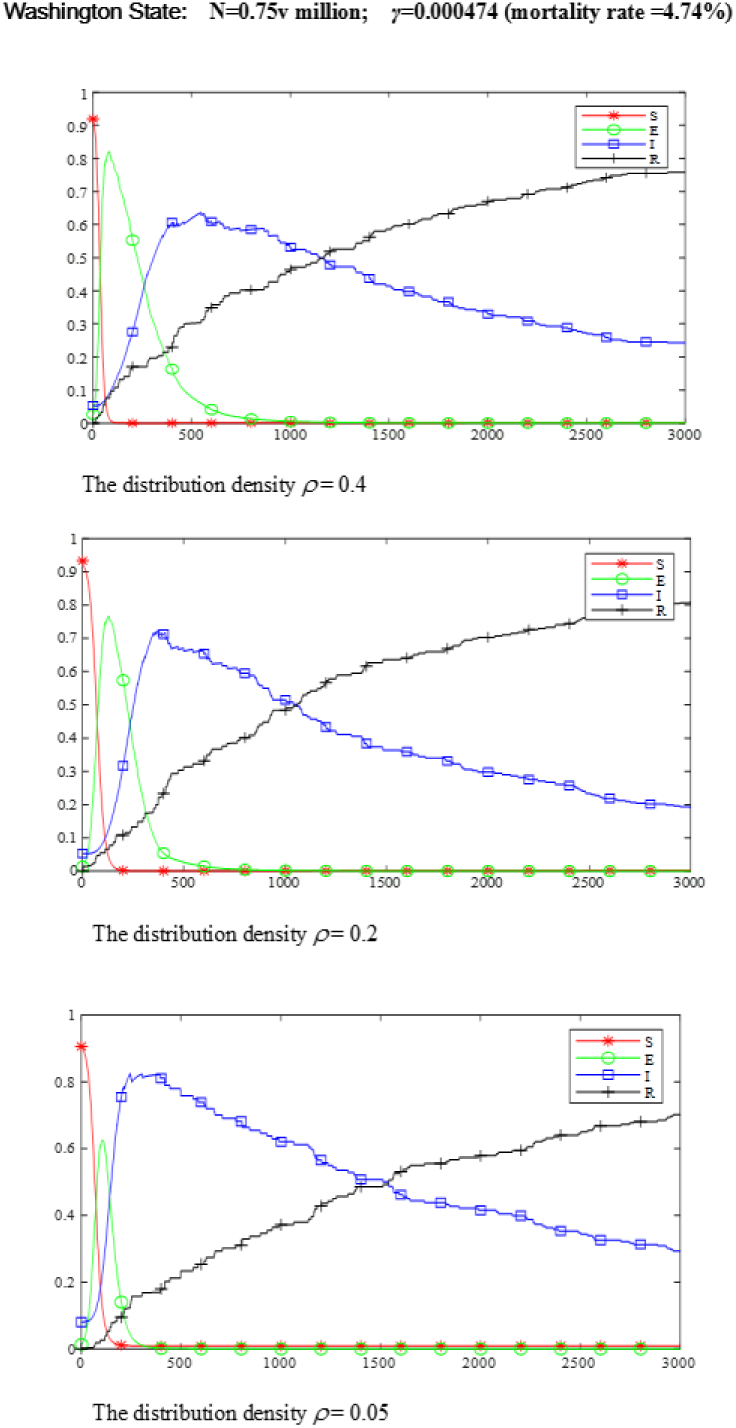

**Figure 5:**
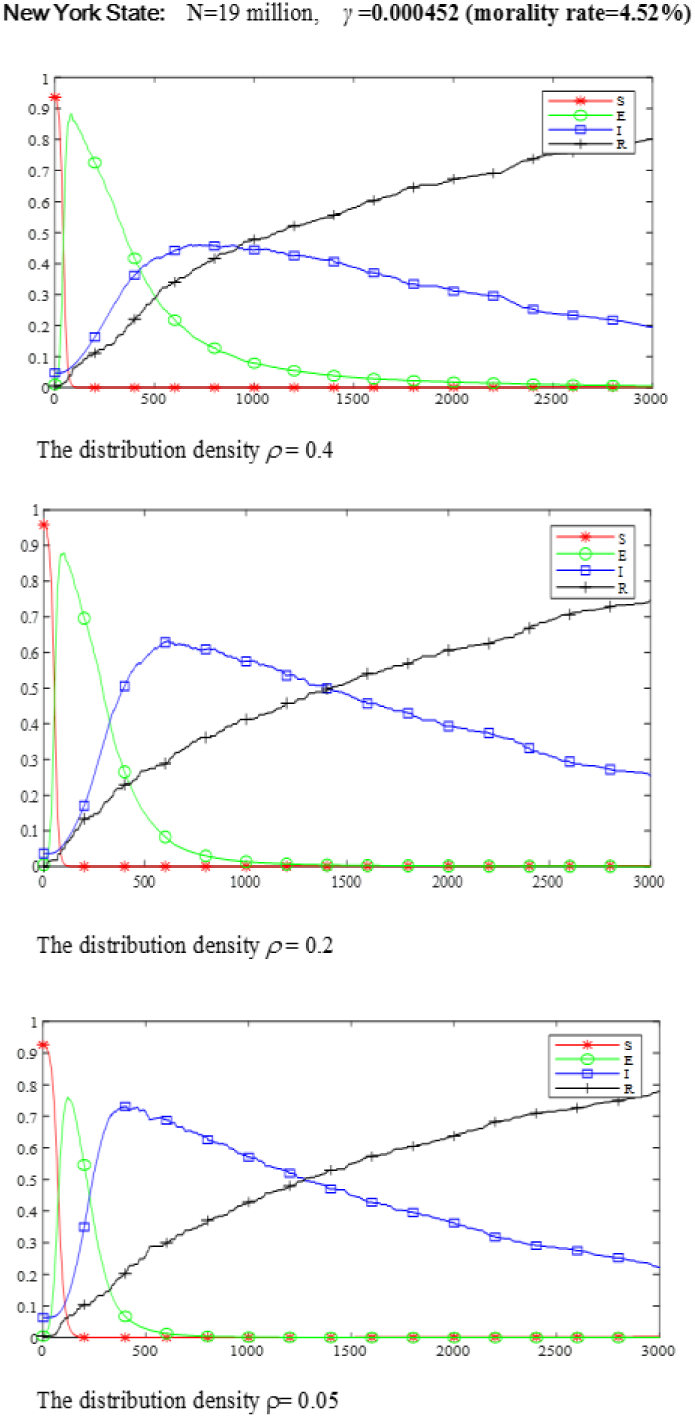

**Figure 6:**
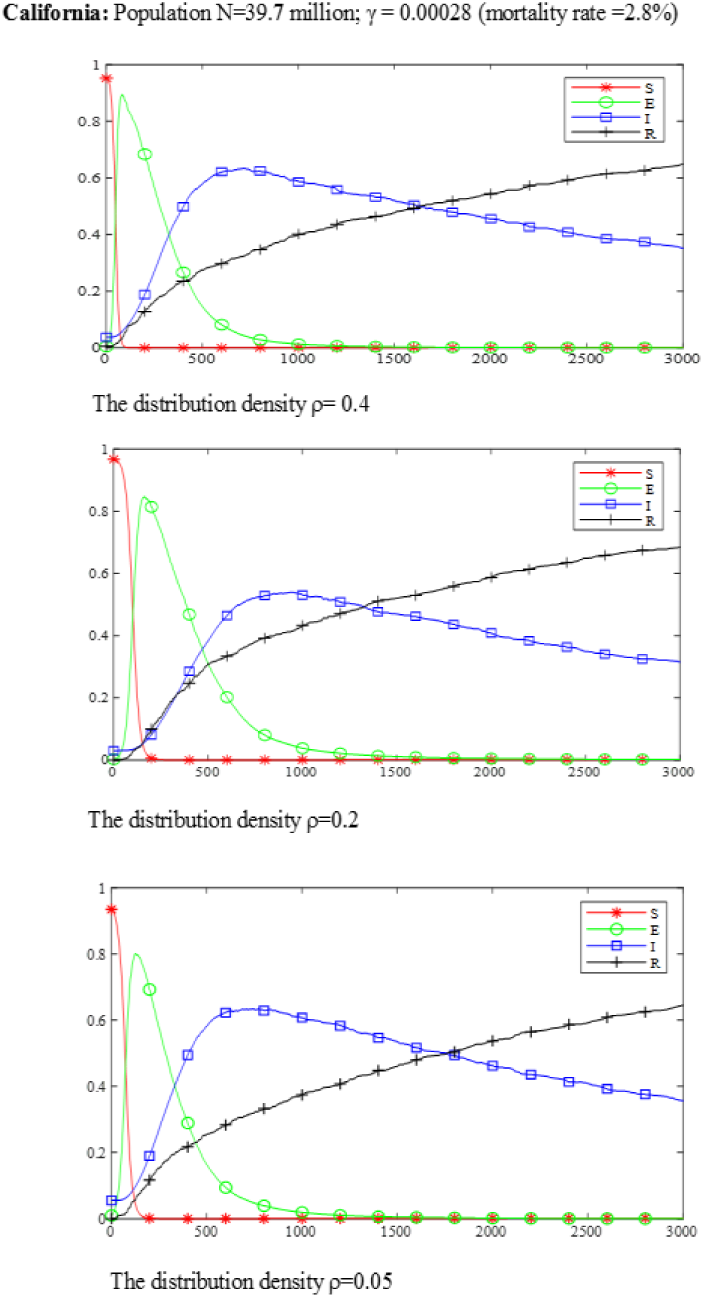

**Figure 7:**
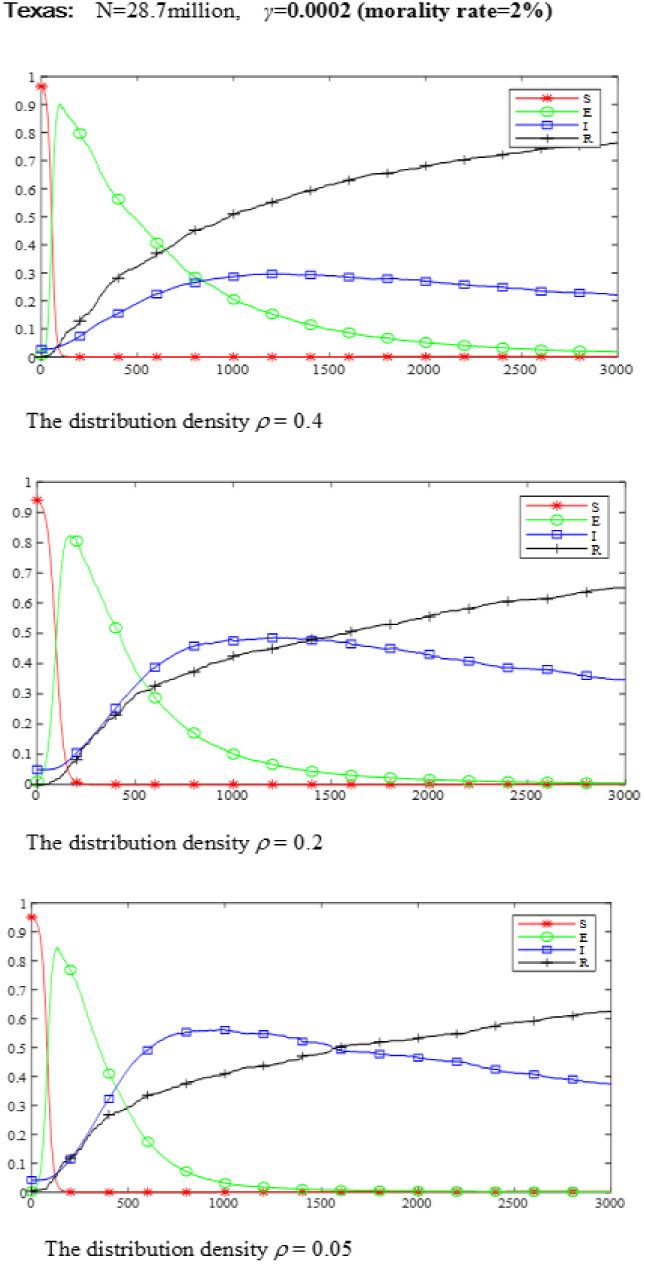

First, based on Figures 4 and 5 below, with numerical results from simulations as of April 10, 2020, we have that

1. For Washington state, both E and I are starting to turn done in around 550 steps (which is around 3.8 days away, as each unit is 10 minutes), thus **around April 14, 2020**, this state is entering the stage that outbreak of COVID-19 is under the control.
2. For New work state, as both E and I are starting to turn done in around 600 steps (which is more than 4 days away), thus **around April 15, 2020**, this state is entering the stage that the outbreak of COVID-19 is under the control.

Second, based on Figures 6 and 7 below, with numerical results from simulations as of April 10, 2020, we have that

1. For California, both E and I are starting to turn done in around 800 steps (which is 6 days away), thus **around April 16, 2020**, this state is entering the stage that the outbreak of COVID-19 is under the control.
2. For Texas, both E and I are starting to turn done in around 1000 steps (which is 7 days away), thus **around April 17, 2020**, this state is entering the stage that the outbreak of COVID-19 is under the control.

By putting all simulations results for 15 Sates together under the framework of iSEIR model, we have the following general conclusion for the outlook by classifying them into four categories as follows.

**Category 1:** Staring **around April 14, 20202**, three states which are **Washington State, Louisiana** and **Indiana** are entering the stage that the outbreak of COVID-19 is under the control, which means daily change of new patients is less than 10% and the gamma is less than zero in general.

**Category 2:** Staring **<u>around</u> <u>April</u> <u>15</u>, 20202**, two states which are **New Jersey, and New Yor**k are entering the stage that the outbreak of COVID-19 is under the control, which means daily change of new patients is less than 10% and the gamma is less than zero in general.

**Category 3:** Staring **<u>around</u> <u>April</u> <u>16</u>, 20202**, seven states which are **California, Florida, Georgia (GA), Illinois, Maryland, Indiana, Michigan**, and **Pennsylvania** are entering the stage that the outbreak of COVID-19 is under the control, which means daily change of new patients is less than 10% and the gamma is less than zero in general.

**Category 4:** Staring **<u>around</u> <u>April</u> <u>17</u>, 20202**, three states which are **Texas, Massachusetts**, and **Connecticut** are entering the stage that the outbreak of COVID-19 is under the control, which means daily change of new patients is less than 10% and the gamma is less than zero in general.

By putting all analysis above together, we are able to classify 15 States into four categories by the different time intervals with the state that the outbreak of COVID-19 is under our control with the criteria we settle that “daily change of new patients is less than 10% and the gamma is less than zero in general” as discussed above.

Our analysis above also shows that by using the concept of “Turning Phase” with the identification of the starting time for the turning phase through the simulation for the observation of “supersaturation phenomenon”, which allows us to conduce different levels for the battle with Outbreak of COVID-19 ongoing base. But we do strongly emphasize that the framework for the prediction of different phases by using iSEIR model accounts for intervention policies and methods such as isolation control programs (e.g. quarantine, implemented in February 2020 in China (see [33] -[34]). Beyond using our iSEIR model for the study of the outbreak of COVID-19 in China from late December to early March 2020, we hope to further apply it to outbreaks worldwide as the study we conducted in this report.

Before arriving at our final remarks, we want to consider the following with regards to the current global state of the COVID-19 outbreak:

1. The virus can be spread by infected and asymptomatic individuals;
2. At current, there is no fully effective medicine or treatment; and
3. No vaccine.

With these conditions, it is important that our iSEIR model be used to aid in modeling the timeline of the COVID-19 outbreak in other countries in shaping public policy. However, we acknowledge that our model is patterned to reflect effective intervention methods and protocols such as the “Wuhan Quarantine” (see [36]). In conclusion, the simulated timeline provided by our iSEIR model best fits outbreak scenarios where early adoption of public health controls and restrictions are implemented to flatten the curve.

Moreover, we want to reinforce that emergency risk management is always associated with the implementation of an emergency plan. The identification of the Turning Time Period is key to emergency planning as it provides a timeline for effective actions and solutions to combat a pandemic by reducing as much unexpected risk as soon as possible. We can further improve our ability to emergency-plan in urgent events, such as in the control of an infectious disease outbreak like COVID-19 in three key areas:

1. a better spatiotemporal model to describe the mechanics of the spread of infectious diseases;
2. an efficient way to conduct numerical simulation to identify the “Turning Time Period (Phase)” for the emergency event, e.g., the timeframe for the outbreak of infectious diseases spread such as OVID-19 virus; and
3. carrying out effective predictive analysis by establishing a coherent bigdata method for data fusion from different sources with different structures to support a dynamic management to respond to daily issues with effective emergency planning.

Finally, like the one did by this paper, taking the data of April 9, 2020, we are able to classify 13 countries in Europe into the following three categories for the outlook of their battle with COVID-19 currently (see Yuan et al. [39] for more in details) stated as follows:

1. Nine countries (consisting of Germany, France, Italy, Spain, Belgium, Austria, Switzerland, Netherlands and Portugal) are in the stage for the control of Outbreak of COVID-19 from April 9, 2020 in less than one week, which means the turning point is appearing;
2. UK is going into the control stage of COVID-19 in two weeks (around April 23, 2020), and Norway and Sweden are in between nine countries and UK, which means their turning points are ahead of UK’s one; and
3. We do not know when Russia is going into the stage to approaching the control level for the outbreak of COVID-19 in next more than weeks to the time before the end of April, 2020.

## Data Availability

No data special for the paper.

## Appendix 1: The Framework of our iSEIR Dynamic Model with Multiplex Networks

For the convenience of our discussion, we give an introduction on the general framework of our iSEIR model which was introduced in [28] (see also [38]) for more in details and numerical simulations lined to the applications.

In brief, our iSEIR model operates under a probability perspective for each individual with the name “**individual Susceptible-Exposed-Infective-Removed (iSEIR)**”, which is an extension of the classic SEIR one. The iSEIR model allows us to conduct simulations from the individual levels located on the nodes of different community networks by incorporating its uncertainty with the probability to conduct random simulation in the corresponding multiplex network.

### 1.1 The Classical SEIR Model

For the SEIR model, the state S refers to the susceptible group (or ignorants) who are susceptible to the disease but have not been infected yet; state E refers to the exposed group who are infected but are not infectious yet; state I refers to those infected who also become infectious; and state R refers to those who have recovered from the infection (through treatment or natural recovery) and are no longer infectious. We also use S(t), E(t), I(t) and R(t) to represent the proportion of the population being in state S, E, I and R at time t, respectively. In the present case, the system of ODEs describing the dynamics of an SEIR epidemic model thus has the following form:

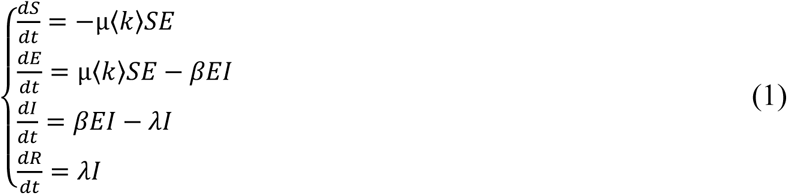

where µ is the rate at which an exposed individual becomes infective, λ is the recovery rate, and with normalization condition (each variable is in percentage change)

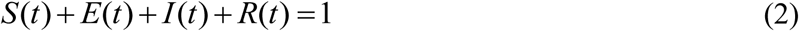

for every t ≥ 0 (since the population is considered closed).

The above deterministic SEIR model and its generalizations have received a lot of attention from various researchers. Indeed, the SEIR model represents more accurately the spread of an epidemic than the corresponding SIR model that does not take into account the latent period. The SEIR model has a slower growth rate, since after the pathogen invasion the susceptible individuals need to pass through the exposed class before they can contribute to the transmission process, as shown (by Figure I_SEIR model illustration) below:

**Figure I.**
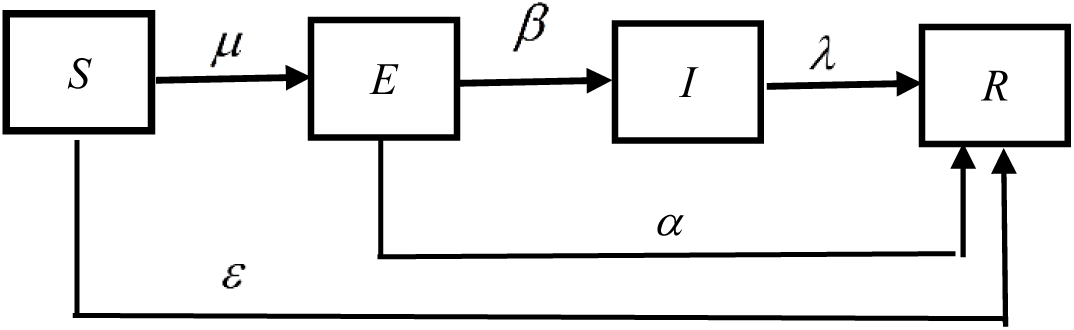
_SEIR model-Illustration.

### 1.2 The Framework of Our iSEIR Dynamic Systems with Multiplex Networks

Based on the former discussion, the iSEIR (see Figure_II_iSEIR model Illustration below) is an extension of SEIR model, but presented in different expression, which is a component form as follows in Eq.(3).

**Figure II.**
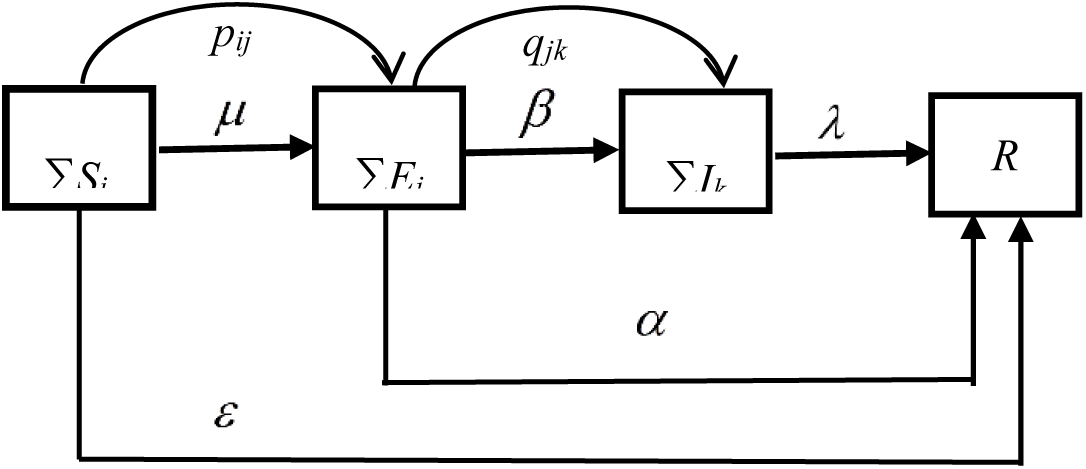
_iSEIR model Illustration.

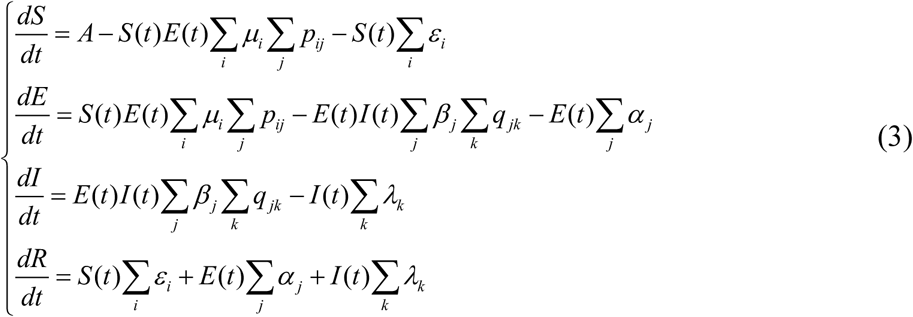

where, parameters in the systems are illustrated as following:

- where A is the growth rate of new arrivals.
- μi denotes the transfer probability from S(t) to E(t).
- pij denotes the connection probability of the i-th sample in S(t) to the j-th sample in E(t); it equal to 1 if connected or 0 if not.
- εi denotes the transfer probability from S(t) to R(t), which is the removed probability.
- βj denotes the transfer probability from E(t) to I(t).
- qjk denotes the connection probability of the j-th sample in E(t) to the k-th sample in I(t); it equal to 1 if connected or 0 if not.
- αj denotes the transfer probability from E(t) to R(t).
- λk denotes the transfer probability from I(t) to R(t).

The proposed iSEIR model is shown as in Figure II_iSEIR model Illustration below:

In order to make the system (3) short and analyze easily, we use the following denotation:

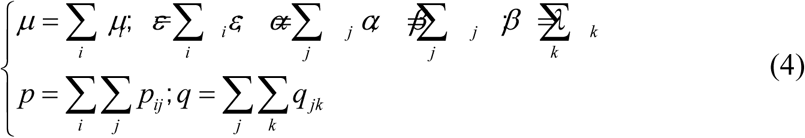

As our iSEIR model is based on the framework of multiplex networks, we also give some notations here, more in details are given in [28] (and also [38]).

By suppose the population S for the spreading consists of N individuals S_j_, j =1, …, N; namely S: = { S_j,_ j = 1, …, N}, and also suppose these N individuals are distributed over M continuous domains Ui, i = 1,……, M, where a domain may refer to a residential district or a network. Then we can conduct the simulation based on framework of probability respective for each individual based on iSEIR in a population multiplex network by following five steps:

**Step 1:** We first allow the transition from state S to state R directly with probability ε per unit time (the same below) by following equation:

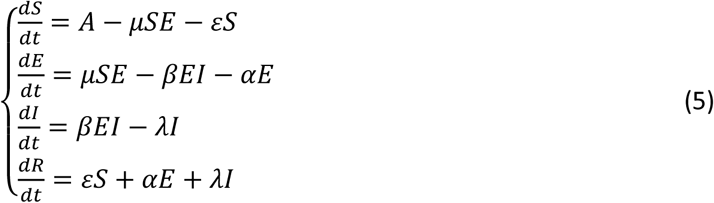

where A is the growth rate or new comer; ε is the probability of a susceptible person being directly transformed into an immune person by means of e.g., isolation; µ is the rate of a susceptible being infected; β is the rate of an infected person becoming infectious; α is the rate of an infected person becoming immune directly; and λ is the rate of an infectious person entering into an immune state. Figure 1 below gives a visual presentation of (5). Note that there is no direction transition between S and I in (5) as illustrated by Figure 1 above.

Here we like to share that the discretization format of iSERI model (5) can be done by using common Euler Difference method.

**Step 2:** Each individual in the rumor spreading network at time t is identified by its state and position in that state group.

**Step 3:** We will establish an adjacency matrix to describe the influence effects between individuals.

**Step 4:** Computing the probabilities of transitions between states involves considering the following two aspects (the K-adjacency method).

**Step 4**.**1:** the distances between uninfected individuals and their neighborhoods of infected individuals within; and

**Step 4**.**2:** the number of individuals infected.

Here we like to point out that the probability of state transition is calculated by k-adjacency method, referring to two factors: (1) the distance between uninfected and infected individuals in the neighborhood; and (2) the number of infected individuals; The use of the K-adjacency method: use the adjacency matrix of n * n to calculate the distance between each individual and other N-1 individuals.

**Step 5:** The full specification of the model is given by combining steps 1 to 4 together with an individual-level representation of (5) that is illustrated in Figure 2 above.

These five steps will help us to run the simulations for the observation of the so-called “Supersaturation Phenomenon” under the probability perspective of all individuals by applying iSEIR mode which help us to identify the “Turning Phase”, which are critical for any emergency plan being successful by responding to the challenge any outbreak of pandemics under the emergency in the practice. Thus the iSEIR will be used as a tool for us to discuss how we can establish the framework for the prediction of the critical “Turning Phase” for the emergency implementation response in an epidemic infectious disease outbreak described in next section.

Finally, the specifications of inputs for related parameters and assumptions are given by the **Appendix 2** below.

## Appendix 2: The General Inputs for the Simulation of iSEIR Model

| The General Inputs for the Simulation under the framework of iSEIR Model |  |
| --- | --- |
| Date: The actual observation from a given day.<br>The data in highlight with yellow color means on the true real scene for the simulation |  |
| Name of Parameter | The specification for each parameters' values |
| # of simulations | 100 times of simulations (unless specified) |
| $N$: # of groups/communities | Specified in simulation, |
| $M$: # of cities | 10 (the value to use unless specified other values) |
| $\rho$: distribution density | 0.4 (the value to use unless specified other values) |
| $c$: distance | base on Euclidean distance formula for each two samples |
| $T$: time steps | 1~3000 (one unit is around 10 minutes unless specified). |
| $N$: population | The true population input based on the true real scene for the simulation by given regions/counties. |
| $i, j, k$: random individual | Uniform distribution |
| $i, j, k$: | The range is from $N$ specified by true real scene |
| $\gamma$ | The true Mortality ratio as input observed from true real scene for the simulation by a given date |
| $A$ | = 0.000001 |
| $p_{ij}$ | = 1, if the distance of $i$ and $j < c$;<br>= 0, otherwise. |
| $q_{jk}$ | = 1, if the distance of $j$ and $k < c$;<br>= 0, otherwise. |
| $\mu_i$ | = 1, if in [0.0001, 1];<br>= 0, otherwise. |
| $\varepsilon_i$ | = 1, if $\leq 0.0001$;<br>= 0, otherwise. |
| $\beta_j$ | = 1, if in [0.001, 1];<br>= 0, otherwise. |
| $\alpha_j$ | = 1, if $\leq 0.001$;<br>= 0, otherwise |
| $\lambda_k$ | = 1, if ≤ the specified value for a given input parameters ($\gamma$)<br>= 0, otherwise |
| Other parameters | Based on the situation to specify (if necessary) |

## Notes

### Competing Interest Statement

The authors have declared no competing interest.

## References

[1] X. Liu, Geoffrey Hewings, Shouyang Wang, Minghui Qin, Xin Xiang, Shan Zheng, Xuefeng Li. Modeling the situation of COVID-19 and effects of different containment strategies in China with dynamic differential equations and parameters estimation. medRxiv preprint doi: https://doi.org/10.1101/2020.03.09.20033498, 2020.

[2] J. L. Murray. Forecasting COVID-19 impact on hospital bed-days, ICU-days, ventilator days and deaths by US state in the next 4 months. MedRxiv. 26 March 2020. doi:10.1101/2020.03.27.20043752.

[3] Joseph T. Wu, Kathy Leung, Mary Bushman, Nishant Kishore, Rene Niehus, Pablo M. de Salazar, Benjamin J. Cowling, Marc Lipsitch & Gabriel M. Leung. Estimating clinical severity of COVID-19 from the transmission dynamics in Wuhan, China. Nature Medicine (2020), 19 March 2020. https://doi.org/10.1038/s41591-020-0822-7.

[4] J. T. Wu, Leung, K. & Leung, G. M. Nowcasting and forecasting the potential domestic and international spread of the 2019-nCoV outbreak originating in Wuhan, China: a modelling study. Lancet 2020; 395: 689–97. https://doi.org/10.1016/S0140-6736(20)30260-9.

[5] Kiesha Prem, Yang Liu, Timothy W Russell, Adam J Kucharski, Rosalind M Eggo, Nicholas Davies. The effect of control strategies to reduce social mixing on outcomes of the COVID-19 epidemic in Wuhan, China: a modelling study. Lancet Public Health 2020. Published Online March 25, 2020. https://www.thelancet.com/journals/lanpub/article/PIIS2468-2667(20)30072-4/fulltext.

[6] Cuilian Li, Chen Li Jia, Chen Xueyu, Zhang Mingzhi, Pang Chi Pui, Chen Haoyu. Retrospective analysis of the possibility of predicting the COVID-19 outbreak from Internet searches and social media data, China, 2020. Euro Surveill. 2020;25(10):pii=2000199. https://doi.org/10.2807/1560-7917.ES.2020.25.10.2000199

[7] Qianying Lin, Shi Zhao, Daozhou Gao, Yijun Lou, Shu Yang, Salihu S. Musa, Maggie H. Wang, Yongli Cai, Weiming Wang, Lin Yang, Daihai He. A conceptual model for the coronavirus disease 2019 (COVID-19) outbreak in Wuhan, China with individual reaction and governmental action. International Journal of Infectious Diseases (93) (2020), 211–216.

[8] T. Kuniya. Prediction of the Epidemic Peak of Coronavirus Disease in Japan, 2020. J. Clin. Med. 2020, 9, 789; doi:10.3390/jcm9030789.

[9] K. Roosa, Y. Lee, R. Luo, A. Kirpich, R. Rothenberg, J.M. Hyman, P. Yan, G. Chowell. Real-time forecasts of the COVID-19 epidemic in China from February 5th to February 24th, 2020. Infectious Disease Modelling, 5(2020), 256–263.

[10] Z. Cao, Zhang Q, Lu X, et al. Estimating the effective reproduction number of the 2019-nCoV in China. medRxiv 2020(a); doi: https://doi.org/10.1101/2020.01.27.20018952.

[11] Z. Cao, Zhang Q, Lu X, et al. Incorporating human movement data to improve epidemiological estimates for 2019-nCoV.MedRxiv 2020. doi: https://doi.org/10.1101/2020.02.07.20021071.

[12] B. J. Cowling and G.M. Leung. Epidemiological research priorities for public health control of the ongoing global novel coronavirus (2019-nCoV) outbreak. Eurosurveillance 2020. doi: https://doi.org/10.2807/1560-7917.ES.2020.25.6.2000110.

[13] S.W. Hermanowicz. Forecasting the Wuhan coronavirus (2019-nCoV) epidemics using a simple (simplistic) model. medRxiv 2020; doi: https://doi.org/10.1101/2020.02.04.20020461

[14] Q. Li, Guan X, Wu P, et al. Early transmission dynamics in Wuhan, China, of Novel Coronavirus–infected pneumonia. New England Journal of Medicine 2020(a); doi:10.1056/NEJMoa2001316.

[15] W. J. Guan, Ni Z Y, Hu Y, et al. Clinical characteristics of 2019 novel coronavirus infection in China. New England Journal of Medicine 2020, doi: 10.1056/NEJMoa2002032.

[16] Norden E. Huang, and Fangli Qiao. A data driven time-dependent transmission rate for tracking an epidemic: a case study of 2019-nCoV. Science Bulletin, 65(2020), 425–427.

[17] Caolin Gu, Jie Zhu, Kai Zhou, Jiang Gu and Yifei Sun. The Infection Point about COVID-19 May Have Passed. Science Bulletin, (2020), to appear, 2020.

[18] Zixin Hu, Qiyang Ge, Shudi Li, Li Jin, and Momiao Xiong. Artificial Intelligence Forecasting of Covid-19 in China. Arxiv.org, 2020. https://arxiv.org/ftp/arxiv/papers/2002/2002.07112.pdf.

[19] Jidi Zhao, Jianguo Jia, Yin Qian, Yuyang Cai. Modeling the COVID-19 outbreak and government control measures based on epidemic dynamics (in Chinese). Systems Engineering-Theory & Practice (in review), 2020.

[20] Y. Yan, Y. Chen, Keji Liu, Xinyue Luo, Boxi Xu, Yu Jiang & Jin Cheng. Modeling and prediction for the trend of outbreak of NCP based on a time-delay dynamic system (in Chinese). Sci Sin Math, 2020, 50: 1–8, doi: 10.1360/SSM-2020-0026.

[21] X. Wang, Sanyi Tang, Yong Chen, Xiaomei Feng, Yanni Xiao & Zongben Xu. When will be the resumption of work in Wuhan and its surrounding areas during COVID-19 epidemic? A data-driven network modeling analysis (in Chinese). Sci Sin Math, 2020, 50: 1–10, doi: 10.1360/SSM-2020-0037.

[22] S. Y. Tang, Tang B, Bragazzi N.L, Fan Xia, Tangjuan Li, Sha He, Pengyu Ren, Xia Wang, Changcheng Xiang, Zhihang peng, jianhong Wu & Yanni Xiao. Analysis of COVID-19 epidemic traced data and stochastic discrete transmission dynamic model (in Chinese). Sci Sin Math, 2020, 50: 1–16, doi: 10.1360/SSM-2020-0053.

[23] Sen-zhong Huang, Zhihang Peng & Zhen Jin. Studies of the strategies for controlling the COVID-19 epidemic in China: Estimation of control efficacy and suggestions for policy makers (in Chinese). Sci Sin Math, 2020, 50: 1–14, doi: 10.1360/SSM2020-0043.

[24] H.J. Cui and T. Hu. Nonlinear regression in COVID-19 forecasting (in Chinese). Sci Sin Math, 2020, 50: 1–12, doi: 10.1360/SSM-2020-0055.

[25] Sir Ronald Ross: 1902 Nobel Laureate in Medicine (http://www.nobelprizes.com/nobel/medicine/1902a.html)

[26] W.O. Kermack and A. G. McKendrick. A Contribution to the Mathematical Theory of Epidemics. Proceedings of the Royal Society Lond. A, 115 (1927), 700–721 (https://doi.org/10.1098/rspa.1927.0118.

[27] Norman T. J. Bailey. The mathematical theory of epidemics. Hafner Publishing Co., New York 1957 viii+194 pp.

[28] George Yuan. The iSEIR model: A dynamic epidemic model for rumor spread in multiplex network with numerical analysis (by George Yuan et al.), Working Paper (Internal Report), Soochow University (Suzhou, China, April/2018) (http://arxiv.org/abs/2003.00144), February, 2020.

[29] George Yuan. “A brief explanation for the concept called ‘ Turning Period’ for COVID-19 in China” (see link: https://mp.weixin.qq.com/s/7OfL3g6z_Bb-R2yl7-GIYg), February 7, 2020.

[30] George Yuan. “Novel coronavirus infection (NCP/COVID-19) epidemic analysis report (February 11th)”, report by Wind Financial Terminal, Feb 12/2020 (see link below: https://news.windin.com/ns/findsnap.php?sourcetype=1&id=487522614&code=C2A7B49D4D25&show=wft&device=android&terminaltype=wft.m&version=6.2.1&share=wechat&from=groupmessage&isappinstalled=0)

[31] George Yuan. “The fight against the new coronavirus entered the second half” (https://mp.weixin.qq.com/s/azAkcR7nzST-dppDHFUZ0g), Feb. 16/2020.

[32] “Report by Members on China COVID-19” (http://www.sesc.org.cn/htm/article/article1199.htm), China Society of Systems Engineering, February 18, 2020.

[33] “China has changed course of COVID-19 outbreak through pragmatic approach: WHO expert said” (http://english.cctv.com/2020/02/26/ARTIGoEEqepxqrFvX8Qvcycn200226.shtml), China Xinhua News Agency, February 26, 2020.

[34] “China-WHO Joint report on COVID-19-February 16-24 (http://www.nhc.gov.cn/jkj/s3578/202002/87fd92510d094e4b9bad597608f5cc2c.shtml.), National Health commission of PRC, February 29, 2020.

[35] Shuan Tan. “Feature: The Modern-Day Nostradamus: George Yuan. Asian Pacific Biotech News.” Vol. 24 (No.03) (2020), 32–37 (https://www.asiabiotech.com/24/2403/24030032x.html).

[36] Sharon Begley. “Once widely criticized, the Wuhan quarantine bought the world time to prepare for COVID-19.” Reporting from the frontiers of health and medicine, STATS (https://www.statnews.com/), February 21, 2020 (see the link: https://www.statnews.com/2020/02/21/coronavirus-wuhan-quarantine-bought-world-time-to-prepare/).

[37] John Hull. Options Futures and Other Derivatives (10th ed). Pearson Education. Inc, 2018.

[38] George Yuan, L. Di, Y. Gu, G. Qian, and X. Qian. The Framework for the Prediction of the Critical Turning Period for Outbreak of COVID-19 Spread in China based on the iSEIR Model. April 11, 2020. In medRxiv preprint doi: https://doi.org/10.1101/2020.04.05.20054346.

[39] George Yuan, L. Di, Y. Gu, G. Qian, and X. Qian. The Prediction for the Outbreak of COVID-19 in European Countries by Using Turning Phase Concepts as of April 9, 2020. Working Paper, Soochow University, April 13, 2020.

